# Comparative assessment of SARS-CoV-2 serology in healthcare workers with Abbott Architect, Roche Elecsys and The Binding site ELISA immunoassays

**DOI:** 10.1101/2021.03.19.21253518

**Authors:** Dinesh Mohanraj, Alison Whitelegg, Kelly Bicknell, Malini Bhole, Lorna Taylor, Caroline Webber

## Abstract

**Purpose:** SARS-CoV-2 serology testing is key for assessing seroprevalence and antibody response post-vaccination in immunocompromised patients. Evaluation of current SARS-CoV-2 serological assays have been performed on samples from severe COVID-19 hospitalised patients. However, robust assay development requires assessment in asymptomatic and non-hospitalised individuals to determine if serological assays are sensitive to detect waning and mild antibody responses. Our study evaluated the performance characteristics between two high-throughput SARS-CoV-2 IgG nucleocapsid assays (Abbott and Roche) and The binding site (TBS) Anti-Spike IgG/A/M ELISA kit in healthcare workers.

**Methods:** 236 samples were collected from Portsmouth Hospital University NHS Trust (PHU) and The Dudley Group NHS Trust and analysed for SARS-CoV-2 serology. We derived concordance, agreement and assay performance as well as using receiver operating characteristic (ROC) curves to redefine the assay threshold of the Abbott assay.

**Results:** Result concordance between the Abbott and TBS was 66%. Discrepant samples were analysed using the Roche assay which showed 100% agreement with the TBS assay. In samples analysed >58 days post-PCR, the sensitivity of Abbott and Roche was 100%. In samples analysed >100 days post-PCR the sensitivity of the Abbott assay dropped to 77.2% but remained at 100% for the Roche assay. A redefined Abbott threshold of 0.64 increased the sensitivity to 90% giving results similar to Roche and TBS assays

**Conclusion:** This study demonstrated Abbott assay had a lower sensitivity in comparison to TBS and Roche. Furthermore, TBS can be implemented as a viable alternative for SARS-CoV-2 serology testing where high-throughput assays are not available on site.

Trial registration number and date of registration

Not applicable.

Trial registration number, date of registration followed by “retrospectively registered”

Not applicable.

**Abstract:** Severe acute respiratory syndrome coronavirus 2 (SARS-CoV-2) serology testing is key for assessing seroprevalence and antibody response post-vaccination in immunocompromised patients. Here we performed a comparison between two high-throughput nucleocapsid assays (Abbott SARS-CoV-2 IgG and Roche Elecsys Anti-SARS-CoV-2) and The Binding Site (TBS) anti-Spike IgG/A/M-SARS-CoV-2 ELISA kit. 236 samples were collected across 2 sites, Portsmouth Hospital University NHS Trust (PHU) and The Dudley Group NHS Trust. We derived concordance, agreement and assay performance as well as using receiver operating characteristic (ROC) curves to redefine the assay threshold of the Abbott assay. Result concordance between the Abbott and TBS was 66%. Discrepant samples were analysed using the Roche assay which showed 100% agreement with the TBS assay. In samples analysed >58 days post-PCR, the sensitivity of Abbott and Roche was 100%. In samples analysed >100 days post-PCR the sensitivity of the Abbott assay dropped to 77.2% but remained at 100% for the Roche assay. A redefined Abbott threshold of 0.64 increased the sensitivity to 90% giving results similar to the Roche and TBS assays. In conclusion, this study demonstrated Abbott assay had a lower sensitivity in comparison to TBS and Roche. This study established TBS can be implemented as a viable alternative for SARS-CoV-2 serology testing where high-throughput assays are not available on site. Furthermore, anti-spike assays, such as TBS, could be used to monitor vaccination responses to deduce SARS-CoV-2 population-immunity. Further optimisation studies are required to evaluate the performance characteristics of these assays which could facilitate widescale sero-epidemiological surveillance.

## Introduction

Severe acute respiratory syndrome coronavirus 2 (SARS-CoV-2), the virus responsible for coronavirus disease 2019 (COVID-19), has led to a global pandemic with more than 106 million confirmed infections and 2.3 million fatalities [1]. Since the initial identification of COVID-19, mass testing and case determination have been a mainstay strategy for controlling viral transmission and guiding both public health and political strategies required to mitigate the deleterious consequences of SARS-CoV-2.

SARS-CoV-2 testing is represented by initial detection of the virus in nasopharyngeal specimens which is conducted by real-time PCR (RT-PCR); currently considered as “gold standard” for confirming suspected diagnosis and identifying asymptomatic carriers [2]. Complementing RT-PCR investigations is serological testing, which utilises immunoassays for the detection of SARS-CoV-2 antibodies. Immunoassays either detect specific antibody types (such as IgM or IgG) or total antibody. However, antibody detection usually occurs 5-7 days post-infection, thus, is not representative of acute infection [3]. Whilst degree and duration of immunity conferred by these antibodies are currently unclear, the widescale use for serological testing has been applied at population level, for establishing population exposure. Other uses of serological analysis include assessing individual infection risk and measuring humoral immunity elicited in vaccine trails [4].

In response, several immunoassays have been designed by manufacturers to meet global laboratory infrastructures enabling high-throughput serological analysis. Two of the earliest tests to become commercially available both measure IgG antibodies against the SARS-CoV-2 nucleocapsid protein. The Abbott SARS-CoV-2 IgG assay was initially validated on sera taken from hospitalised COVID-19 patients ≥14 days post RT-PCR (n=31) and 997 pre-pandemic sera, and reported a sensitivity and specificity of 100% (95% CI: 95.89-100.00) and 99.6% (95% CI: 98.98-99.89), respectively [5]. The Roche Elecsys Anti-SARS-CoV-2 assay was initially validated using sera from COVID-19 patients ≥14 days post RT-PCR (n=102) and 10453 pre-pandemic sera, and reported a sensitivity and specificity of 99.5% (95% CI: 97.0-100%) and 99.80% (95% CI: 99.69-99.88%) respectively [6].

The majority of reports on the performance of serological assays have been performed on samples from hospitalised patients with severe COVID-19, who have a high viral load and elicit robust immune responses which may in part account for the high sensitivities reported. To date, there are limited studies which have focused on evaluating assay performance in non-hospitalised or community based COVID-19 cases. From the limited data available in such patients, studies have demonstrated poor sensitivity using the Abbott assay. For instance, a pre-print report highlighted a sensitivity of just 61.5% when investigating community based COVID-19 cases (n=26) [7].

Understanding antibody responses in asymptomatic and non-hospitalised individuals is of major importance for ascertaining viral transmission and for SARS-CoV-2 serological assay development [8]. As a result, it is crucial that antibody assays are able to detect low-levels of antibodies in order to be of use both in clinical and sero-epidemiological settings. The Binding Site (TBS) Anti-IgG/A/M SARS-CoV-2 ELISA (TBS, Birmingham, UK) measures IgG, IgA and IgM antibodies against the SARS-CoV-2 trimeric spike glycoprotein, and has shown to detect antibodies in PCR-confirmed non-hospitalised asymptomatic COVID-19 patients and patients with mild disease [9].

To date, limited direct assessments of multiple immunoassays have been conducted on large data sets from non-hospitalised patients. We therefore sough to evaluate the Abbott, Roche and TBS immunoassays in samples taken from healthcare workers to identify both assay sensitivities and redefine assay thresholds required for optimisation.

## Methods

(must include sufficient information to allow readers to understand the article content)

### Patients and Study Design

To date, the UK has limited established standards for assessment of performance metrics for SARS-CoV-2 immunoassays internationally. There is no specific guidance on assay performance from either the US FDA or the European Centre for Disease Prevention and Control. A head to head comparison of different SARS-Cov-2 immunoassays was conducted at two different sites: Queen Alexandra Hospital, Portsmouth Hospitals University NHS Trust (PHU) and Russell Hall Hospital (RH), Dudley Group NHS Foundation Trust. At PHU, this study was approved by Portsmouth Hospital NHS Trust Research Ethics Committee. Serum samples (n=188) were collected that had been provided as part of the SIREN-research study (Clinicaltrails.gov identifier: NCT12345678). Healthcare workers, aged 18 years or greater provided fortnightly blood samples for SARS-CoV-2 serology surveillance. Only 5 participants had positive SARS-CoV-2 RT-PCR prior to SIREN enrolment. Additionally, 10 presumed COVID-19 negative samples, aged 18 years or greater were included in the study. At RH, serum from healthcare workers (n=64), aged 18 years or greater, was collected, 39 samples were RT-PCR confirmed positive for SARS-CoV-2 and 9 were RT-PCR negative and symptom negative for SARS-CoV-2 [10].

### Anti-SARS-CoV-2 assays

Samples from PHU were initially analysed for SARS-CoV-2 antibodies at the Department of Medical Microbiology, PHU, using the Abbott nucleocapsid IgG assay [Abbott, Chicago, IL, USA] as part of the SIREN study. Samples were identified as positive or negative based on manufacturers cut-off (appendix, p 1). Samples were further evaluated using The Binding Site (TBS) human anti-IgG/A/M SARS-CoV-2 ELISA [The Binding Site, Birmingham, UK]. If discordance was reported between the TBS and Abbott platforms, samples were evaluated using the Elecsys® Anti-SARS-CoV-2 IgG nucleocapsid immunoassay [Roche, Basel, Switzerland] at Poole NHS Trust. Samples from RH were analysed using the Abbott nucleocapsid IgG assay [Abbott, Chicago, IL, USA] and further evaluated using the Elecsys® Anti-SARS-CoV-2 IgG nucleocapsid immunoassay [Roche, Basel, Switzerland]. Assays were conducted in accordance to manufacturers’ standard operating procedures by HCPC registered laboratory staff in laboratories which hold accredited UK Accreditation service status. All assays were conducted with specified controls and calibrants using clinical cut-off thresholds for negative and positive as determined by the manufacturers.

### Procedures

Primary sample tubes were stored at 2-8°C for up to seven days after venepuncture. Selected samples for each run were derived within this seven day period and primary samples were centrifuged for 10 minutes at 3500g. Supernatant (0.5-1ml) was derived and made up into sample aliquots, which were stored in −20°C until they were processed by respective immunoassays. Freeze-thaw cycles were limited to less than three cycles.

### Statistical Analysis

Analyses were performed using Graphpad Prism v9.0.1 (La Jolla, California, USA) and SPSS v27.0 (IBM Corp, Armonk, NY). Fisher’s exact test was used to evaluate categorical data between immunoassays, whereas Chi-squared analysis was used to evaluate categorical data between 3-way method comparison. One-sample t-test was employed to determine the significance of result outcomes produced by each immunoassay. Sensitivity and specificity with exact binomial 95% CI for assays was carried out by Fisher’s exact test. Assessment of agreement between immunoassays was conducted by concordance (percentage) and by Cohen’s Kappa. Receiver operator characteristics (ROC) curves defined trade-offs in assay sensitivity and specificity for Abbott immunoassay.

### Ethical Considerations

Study proposal was evaluated by Portsmouth Hospital NHS Trust Research Ethics Committee (REC), which deemed the study to be a service evaluation. PHU REC provided a decision stating this study did not require Research Ethics or NHS research confirmations. Samples were derived from altruistic healthcare volunteers which was designed for assay verification. All study protocols used in this study adhered and satisfied standards of ethical practice and was compliant with UK regulations.

## Results

### PHU study: Comparison between Abbott and TBS

Comparing antibody levels using the Abbott and TBS assays in 188 serum samples taken from healthcare workers (HCW) found concordant results in 125 samples (66.4%, Figure 1a) demonstrating a fair agreement between assays as measured using Cohen Kappa analysis (1=0.377, 95% CI: 0.27-0.48, SE=0.054). A two-fold decrease in positive antibody detection was seen with the Abbott assay compared to TBS, demonstrating a significant difference in outcomes between assays (Fisher’s exact test, p<0.0001). Of the samples reported as negative using the Abbott assay (n=131), 59 were reported as positive using the TBS assay (p<0.0001, Figure 1b). As seen in Figure 1b, a clear demarcation of positive and negative samples can be seen using the TBS assay. In these 59 samples, the results from the Abbott assays fell within index values of 0.26-1.31 (Fig 1c).

**Figure 1.**
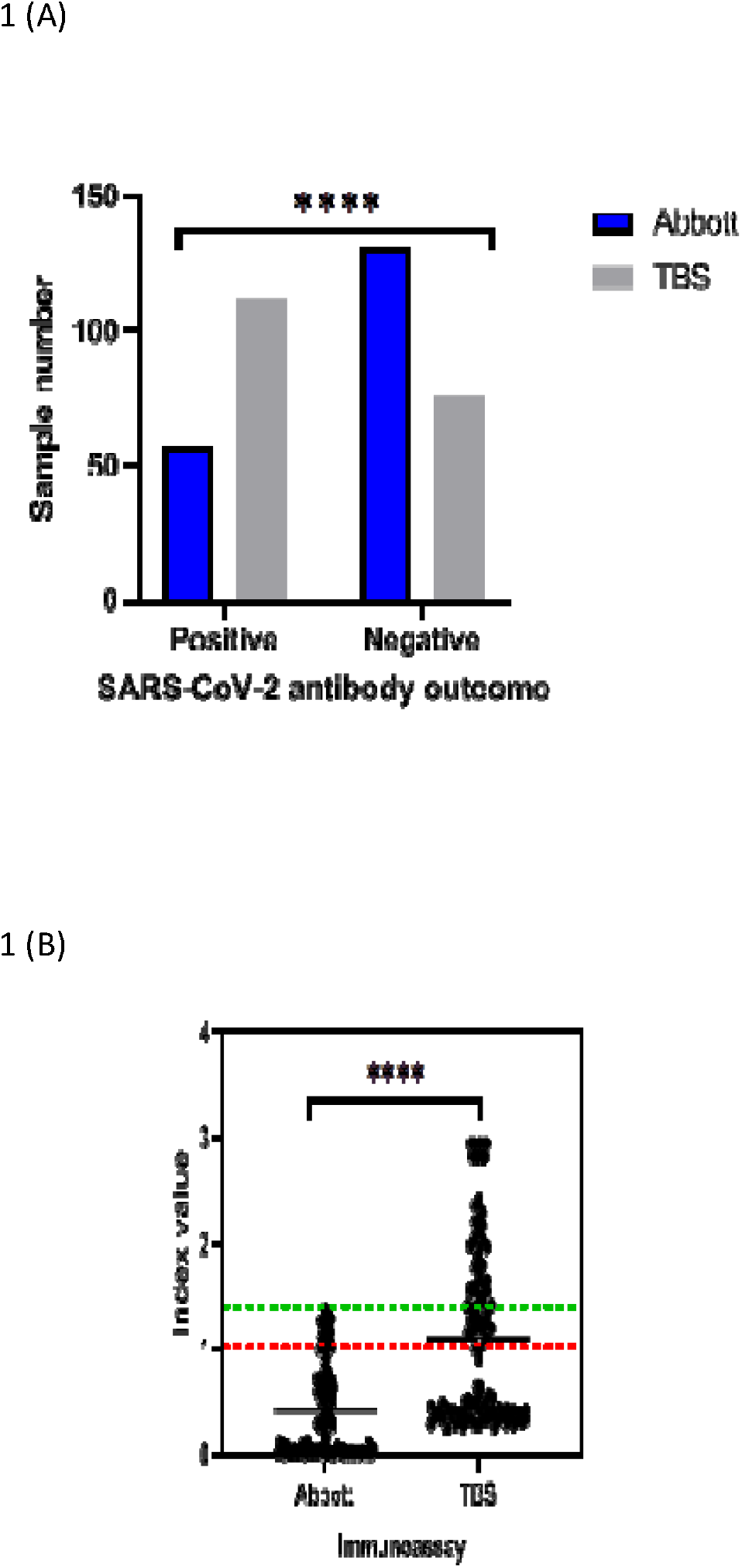
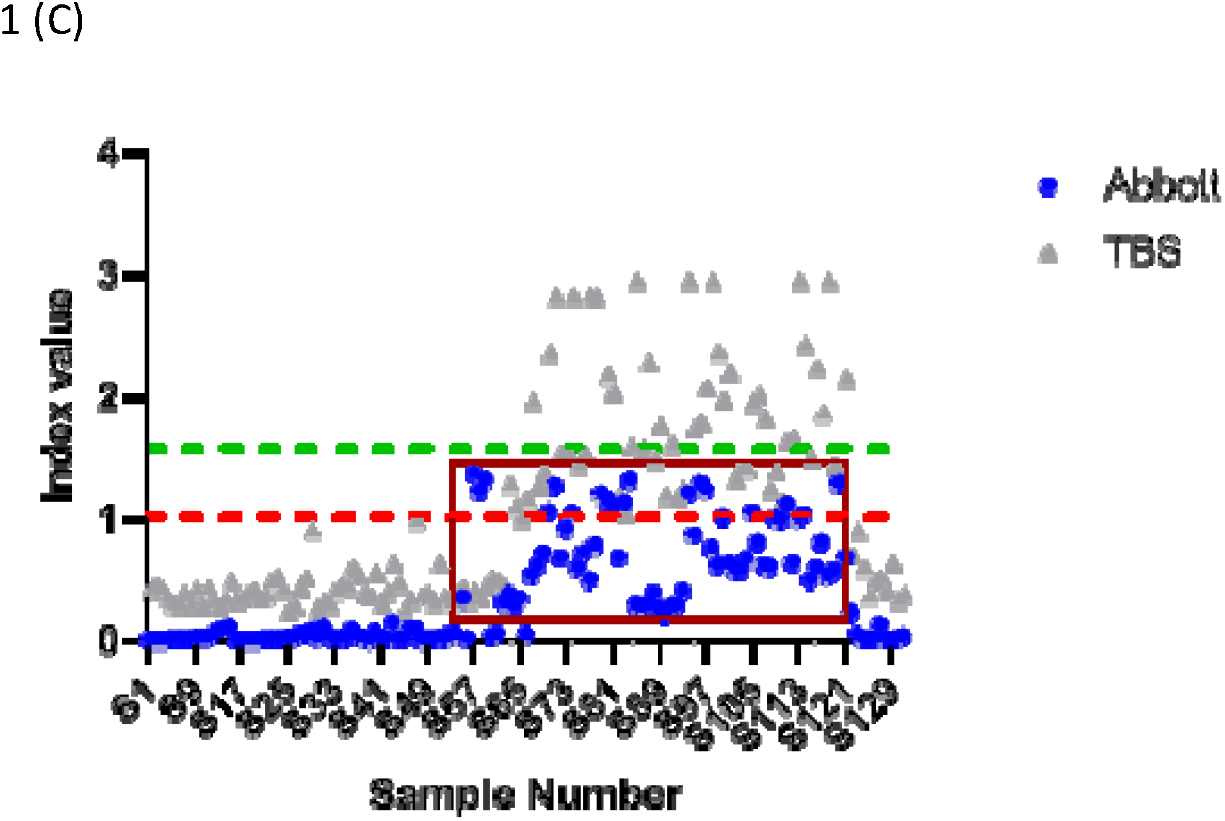
Comparative evaluation between Abbott and TBS. Results are represented as (A) bar graph, which compares positive and negative SARS-CoV-2 antibody outcomes in 188 samples analysed between Abbott and TBS. Statistically significant differences for positive versus negative outcomes derived from both assays (****=p<0.0001, Fisher’s exact test). Nested scatterplot is represented by (B), which portrays the different in outcomes produced by TBS on analysis of 131 samples as deemed negative by Abbott (Green dashed line=cut-off for Abbott (1.40 Index value)). 45% of TBS outcomes were positive (red dashed line= Cut-off for TBS, (1.0 Index value)) and above TBS mean index value (1.06, 95% CI: 0.92-1.20), which was significantly higher compared to TBS (0.36 index value, 95%CI: 0.31-0.46, ****= p<0.0001, one sample t-test). Paired index values derived from negative sample analysis, from Abbott and TBS, are represented on the scatterplot in (C). Discrepant results from Abbott (blue dots) are categorised within the brown rectangular box (n=59, index values; 0.26-1.31), where, reciprocal analysis by TBS (grey triangles) is deemed positive in all 59 samples.

Of the 59 discordant samples, 48 were sent to Poole NHS Trust for analysis using the Roche Elecsys Anti-SARS-CoV-2 IgG nucleocapsid assay. The Roche and Abbott assays were concordant in 2% (1/48) of samples representing poor agreement (⍰ -Cohen= -0.133, 95% CI: −0.26-0.02), whereas concordance between the Roche and TBS assays was 100%. Of the 48 samples tested with all 3 assays, the Abbott assay reported a negative result in 45 samples, whereas the TBS and Roche assays reported negative results in only 4 samples, thus demonstrating a significant difference in outcomes reported by the Abbott versus Roche and TBS assays (Chi-square, p<0.0001, Figure 2).

**Figure 2.**
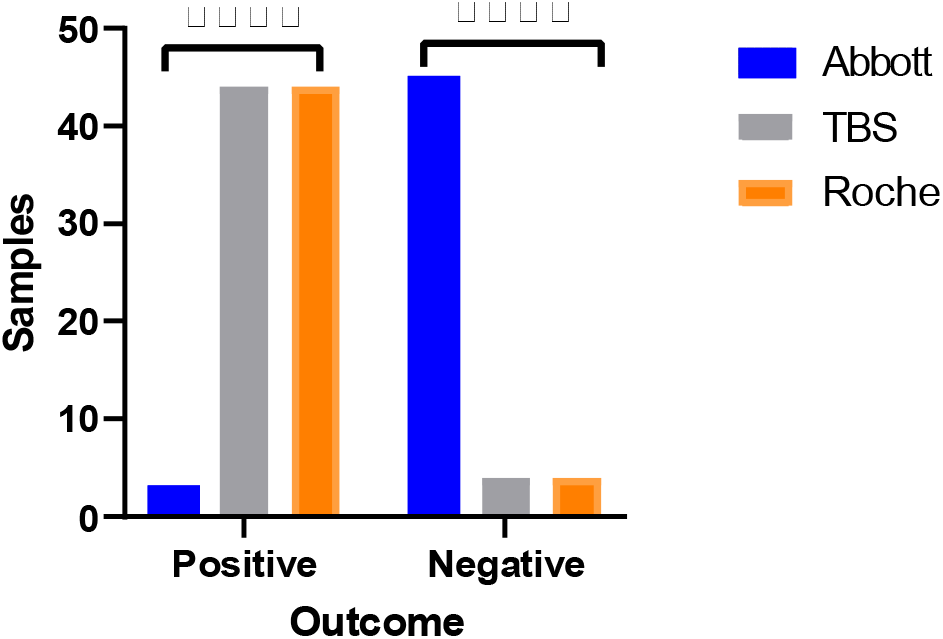
Three-way method comparison between Abbott, TBS and Roche. Bar graph portrays SARS-CoV-2 antibody outcomes produced by Roche in discordant sample-set (n=48) between Abbott and TBS. Statistically significant differences observed from outcomes generated by Abbott versus Roche and TBS (****= p<0.0001, Chi-Square test). Outcomes produced by both TBS and Roche are identical with each other (100% concordance).

### Russell Hall (RH) study; evaluation of sensitivity and specificity between Roche and Abbott

Serum samples were collected from 48 healthcare workers (HCWs) within 58 days of registering a RT-PCR result or onset of COVID-19 symptoms (PCR positive:39, PCR negative:9). A further 16 samples were collected from HCWs who did not have RT-PCR performed and showed no signs and symptoms of COVID-19 infection; classified as presumed COVID-19 negatives. Amongst these samples, 28 were followed up >100 days post PCR-result or symptom onset (PCR positive: 22, PCR-negative: 6).

In samples analysed within ≥19-≤58days, the Abbott and Roche assays showed 100% concordance and perfect agreement (⍰-Cohen: 1.000). All assay characteristics for both the Roche and Abbott assays were identical (Table 1); both assays reported sensitivity and specificity of 97.4% and 80%, respectively, with a LR of 4.87.

**Table 1.** Assay performance characteristics of Roche and Abbott in analysis of HCWs samples both within 58days and >100days of PCR-result or symptom onset. Specificity was assessed using PCR-negative or no-onset of COVID-19 symptoms samples, and sensitivity using RT-PCR positive samples. CI= Confidence interval. SE= Standard error.

| Parameter | Within 58 days post PCR/symptom onset |  | >100 days post PCR/symptom onset |  |
| --- | --- | --- | --- | --- |
|  | Abbott | Roche | Abbott | Roche |
| Sensitivity | 97.4% (95% CI: 86.8-99.8%) |  | 77.3% (95% CI: 56.6- 89.9%) | 100% (95% CI: 85.1- 100%) |
| Specificity | 80.0% (95% CI: 60.9- 91.1%) |  | 83.3% (95% CI: 43.7-99.2%) |  |
| PPV | 0.88 (95% CI: 0.75-0.95) |  | 0.94 (95%CI: 0.74- 0.99) | 0.96 (95%CI: 0.79- 0.99) |
| NPV | 0.95 (95%CI: 0.77-0.99) |  | 0.50 (95% CI: 0.24- 0.76) | 1.00 (0.57-1.00) |
| LR | 4.87 |  | 4.64 | 6.00 |
| κ-Cohen | 1.000 SE: 0.000 | | 0.462 SE: 0.164 | |

Samples analysed after >100days showed a concordance of 79% (22/28) with a moderate agreement (1-Cohen: 0.462, 95% CI: 0.15-0.78). Of the 6 discordant samples, all where reported as negative using the Abbott assay, but positive using the Roche assays. Of these, 5/6 were PCR positive on initial testing, and the sample that was PCR negative tested positive for both assays at the earlier timepoint.

### RH Abbott-Roche comparison consistent with PHU-study discordant range

To further interrogate assay performance, samples that reported low positive results (n=20, range: 1.28-4.00 index value) using the Roche assay were compared to the result generated using the Abbott assay (Figure 3). Concordance was found in only 4/20 samples (20%). Consistent with the findings from PHU, most discrepancies arose in samples that has a result between 0.26-1.31 according to the Abbott assay. This further exemplifies the inferior sensitivity of the Abbott assay, which gave significantly more false negatives (Fisher’s exact test, p<0.0001) in comparison to the Roche assay.

**Figure 3.**
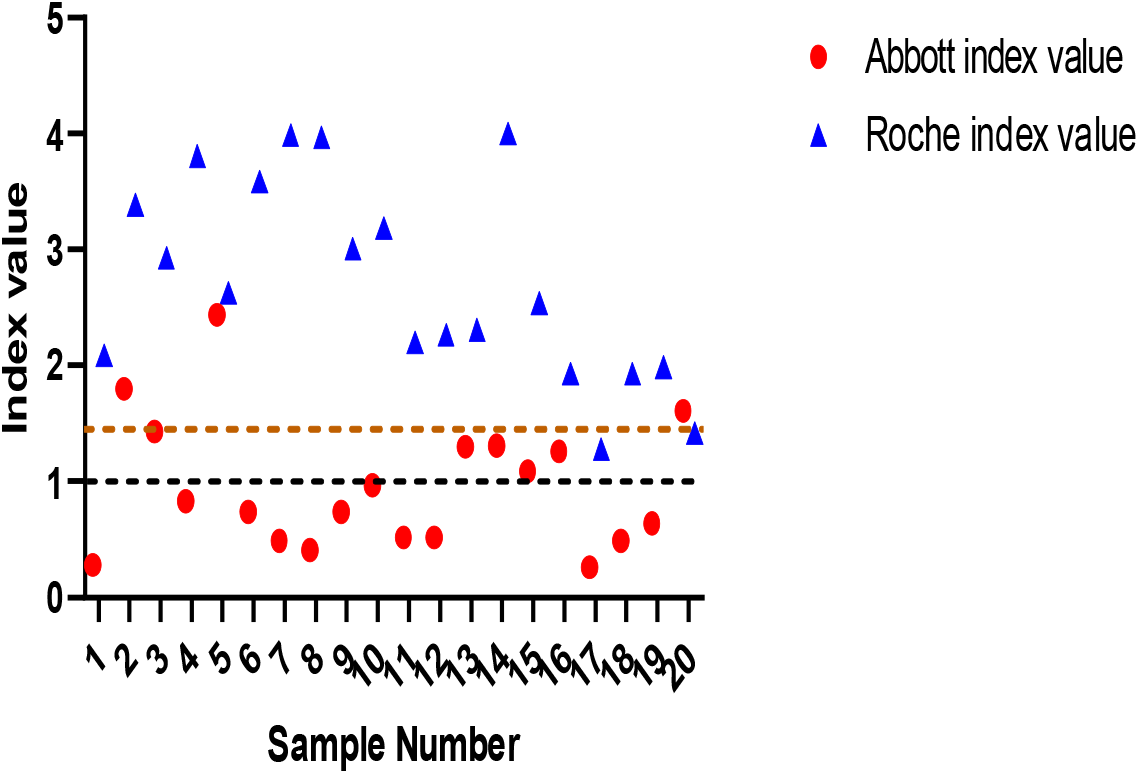
Roche reported low-positive samples analysed by Abbott. Statistically significant difference in reported outcomes observed in low-positive samples analysed by Abbott. All discordant outcomes (n=16) reported by Abbott are below its manufacturers set threshold (brown-dash line) and comprise of index values of 0.26-1.31; consistent with PHU Abbott versus TBS discordant range. Black-dash line= Roche manufacturer threshold. Brown-dashed line= Abbott manufacture threshold.

### Definition of thresholds harmonising Abbott, TBS and Roche results

The above analyses allowed us to define the optimal threshold enabling concordance and agreement between the 3 assays (Figure 4). Based on ROC-analysis, a new threshold of ≥0.64 for the index value of the Abbott assay was selected. Using the ROC-curve, this threshold gave a sensitivity of 90.91% (95% CI: 70.84%-98.88%), specificity of 83.33% (95% CI: 35.88%-99.58%) and LR of 5.45 whereas the manufacturer’s threshold of 1.4 gave of a sensitivity of 77.27% (95%CI: 54.63-92.18%), specificity 83.3% (95%CI: 35.88-99.58) and LR of 4.64.

**Figure 4.**
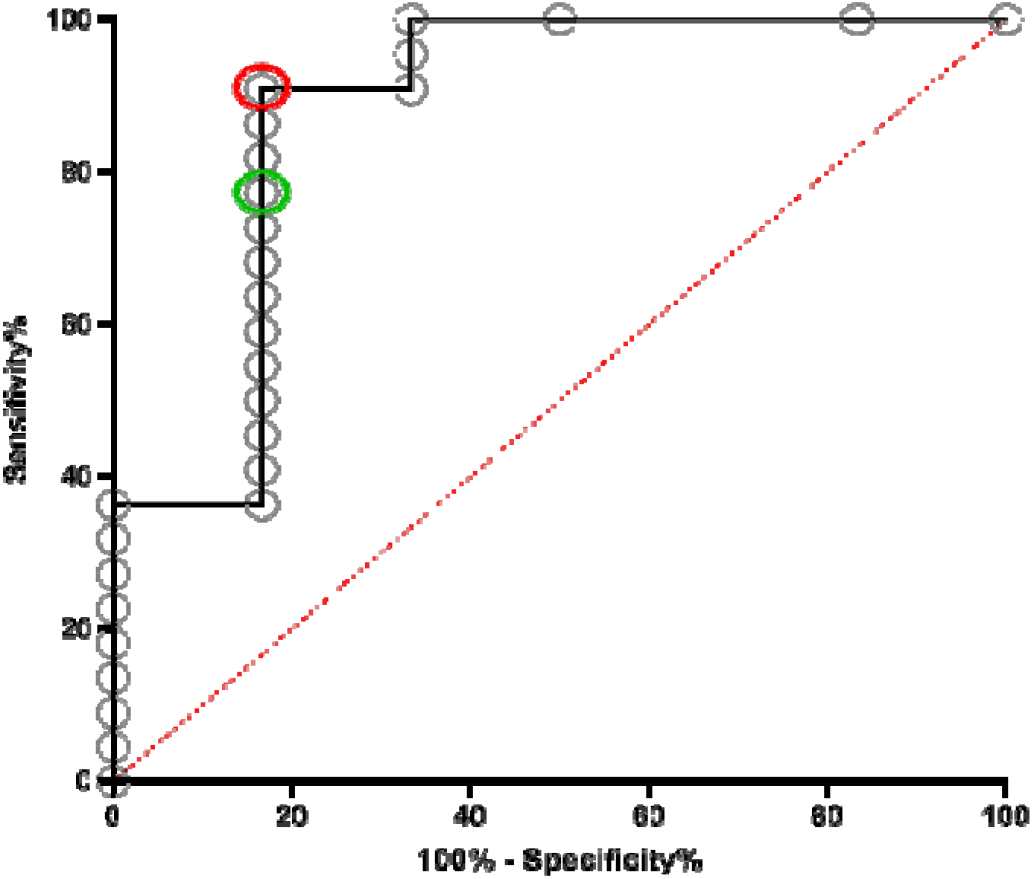
ROC curve for Abbott at specification of samples taken >100 days post-PCR result. The new cut-off (sensitivity= 90.91% (95% CI: 70.84%-98.88%), specificity= 83.33% (95% CI: 35.88%-99.58%)) and manufacturer cut-off (sensitivity=77.27% (95%CI: 54.63-92.18%), specificity= 83.3% (95%CI: 35.88-99.58)) are represented by red and green circles, respectively. Data are presented for 22 RT-PCR-positive samples and 6 known RT-PCR negative samples. Area under curve= 0.878, SE: 0.0991, 95% CI: 0.6845 to 1.000.

Implementation of this redefined threshold to data obtained at PHU (n=188 samples) gave a concordance of 82% (155/188) between the Abbott and TBS assays, where outcomes between assays were not significantly different (Fisher’s exact test, p=0.0621, Figure 5a). This represented an increase in concordance of 16% and resulted in good agreement (K-Cohen: 0.650, 95% CI: 0.54-0.76, SE: 0.054).

**Figure 5.**
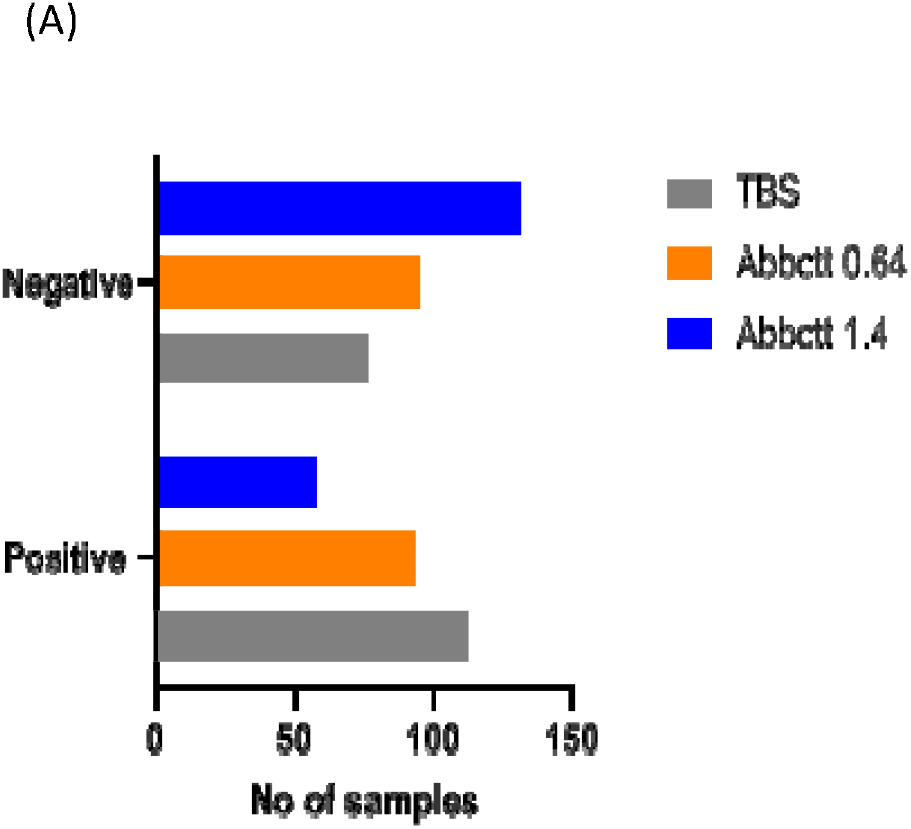
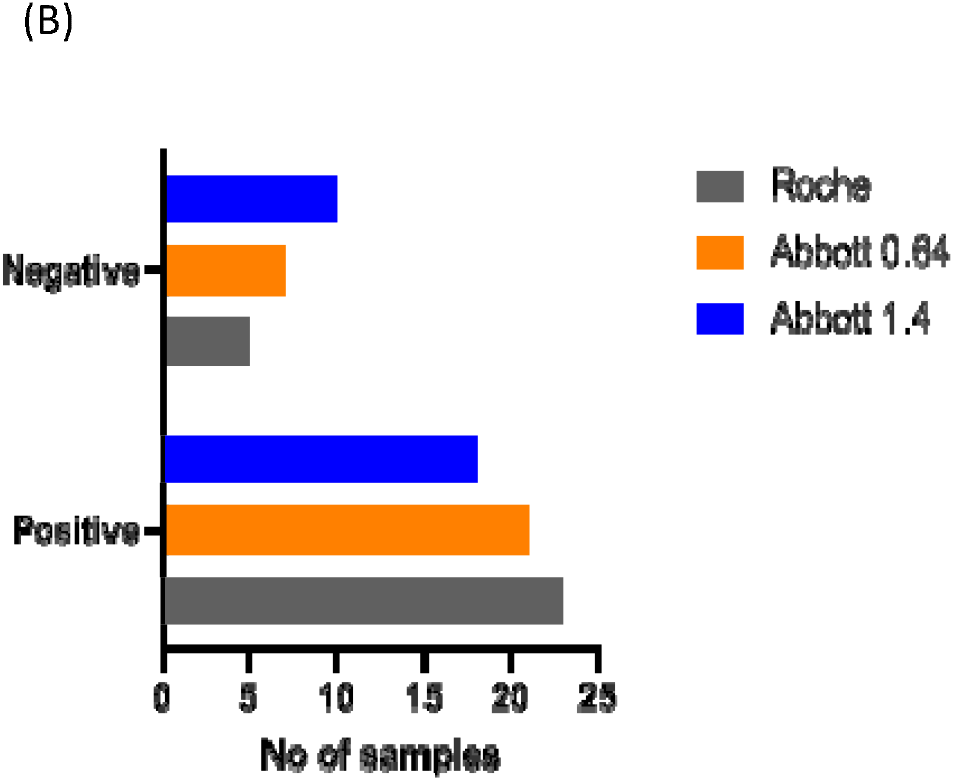
Comparison of SARS-CoV-2 antibody outcomes from use of manufactures Abbott threshold versus redefined Abbott threshold from PHU and RH. No significant differences between antibody outcomes (positive, negative) was observed between Abbott 0.64 (redefined threshold) versus TBS (A) and Roche (B). Both concordance and agreement between Abbott versus Roche and TBS improved from use of Abbott 0.64 threshold compared to use of manufacturers cut-off (Abbott 1.4). Data presented for; (A) comprise of 188 HCWs samples (PHU site) which was initially reported by Abbott and then analysed and reported by TBS, (B) n=28 HCWs samples from RH site, 22 RT-PCR positive and 6 RT-PCR negative samples analysed and reported both on Abbott and Roche.

No significant difference in results between the Roche and Abbott assays was established when the redefined threshold was applied to samples analysed >100 days post-PCR result (Fisher’s exact test, p=0.7458, Figure 5b). Concordance between assays was 89%, which represented an increase of 10% compared to manufacturer’s threshold, and agreement was good between both assays (K-Cohen: 0.650, 95% CI: 0.33-1.00, SE: 0.054). Concordance in low-positive Roche samples (n=20) was improved from 20% to 65%. Together, these findings demonstrate that use of redefined threshold enhances the sensitivity of the Abbott assay in line with that reported by both the Roche and TBS assays.

## Discussion

There is an urgent requirement to identify surveillant strategies to ease national impositions against COVID-19, which will enable the recovery of socio-economic activities. It is known that individuals with neutralising antibodies against SARS-CoV-2 may confer herd immunity [11], thus acting as a barrier against transmission and protecting vulnerable members of society as well as easing the pressure on stretched healthcare services. As a result, serological testing has been implemented at the population level in order provide information on seroprevalence. To date, few direct comparisons of immunoassay performance have been conducted on large sample sets. Moreover, most studies investigating the performance of the Abbott assay have been based on hospitalised patients with severe COVID-19, who have high viral loads stimulating robust antibody responses. Thus, it is not surprising to observe reported claims of sensitivities in the range of 98.3-100% [12-15].

We collected serum samples from healthcare workers (HCW) at two different sites and evaluated the performance of 3 different SARS-CoV-2 antibody assays; TBS, Abbott and Roche. Samples from HCW were stratified according to the data available at each site. At Portsmouth University Hospital (PHU) samples were stratified into positive and negative as reported by the Abbott assay, which were later evaluated using the TBS assay. As samples from HCWs were collected during the first-wave, mass testing of RT-PCR was not underway, thus, most of these participants had no available RT-PCR data. Samples collected at Russells Hall Hospital (RH) were stratified into COVID-19 positive and negative samples according to RT-PCR. At both sites, concordance and agreement of SARS-CoV-2 serology were compared between the different immunoassays.

Significant discrepancies were identified between the Abbott and TBS assays, with only 66% concordance found. Analysis of discrepant samples using the Roche assay showed perfect agreement with the TBS assay, whilst only 1 result concordant between the Roche and Abbott assays. This highlighted the potential of significant underestimation of seroprevalence using the Abbott assay. Of note, the sensitivity and specificity (97.4% and 80.0%, respectively) of the Abbott and Roche assays was identical in samples tested ≤58 days post PCR or symptom onset, but differed significantly in samples tested >100 days post PCR or symptom onset (77.3% by Abbot compares to 100% by Roche). As SARS-CoV-2 antibody levels naturally wane over time [16], this would suggest that the Abbott assay lacks the sensitivity to detect lower antibody levels, which can be detected using the Roche and TBS assays. Our findings also highlight that the manufacturers cut-off threshold ≥1.4 index value, is likely to report false negatives in an assay that already comprises low sensitivity. Uses ROC analysis we defined a cut-off of 0.64 which gave a sensitivity of 90.0% and was showed good agreement with the Roche and TBS assay. Whilst it does not meet MHRA assay stipulation of 98% sensitivity and specificity [19], optimisation of this redefined threshold lead to fewer false negative results.

Our findings are supported by a study that highlighted that 59% of results produced by Abbott were false negatives, which by subsequent analysis using the Virtus (IgM and IgG) test were shown to be positive [17]. Furthermore, the low sensitivity of the Abbott assay was also demonstrated in results from the Spanish national serologic survey [18], which tested 5118 individuals with typical COVID-19 symptoms within 14 days of symptom onset. It was found that only 18.0% of these individuals had SARS-CoV-2 antibodies. The authors interpreted this as “sizable proportion of suspected cases might not have been caused by SARS-CoV-2”. We find this explanation implausible and suggest that this data further supports our consensus that the Abbott assay underestimates population immunity to SARS-CoV-2. It is our opinion that the Abbott assay used in the Spanish study conferred low sensitivity, which is supported by our findings of 77% sensitivity from our data derived from two different hospital sites.

Whilst sensitivity and specificity are crucial parameters for any serological assay, estimating the PPV and NPV is useful when determining clinical and sero-epidemiological utility. Our data demonstrates that Abbott’s low NPV (0.5) is not sufficient to rule out an individual’s recent exposure to SARS-CoV-2, which calls into question the utility of the assay when determining population immunity. We propose that the TBS assay will confer superior clinical and epidemiological utility, compared to the Roche and Abbott assay. As this study demonstrated, the TBS assay demonstrated enhanced sensitivity as it was able to detect waning antibody responses. This was supported by a recent study which demonstrated that the TBS assay was able to detect antibody responses in non-hospitalised asymptomatic COVID-19 patients and patients with mild-disease [9].

The current licenced COVID-19 vaccines target the Spike-protein of SARS-CoV-2, consequently, eliciting humoral responses against Spike-protein. However, Roche and Abbott assays both measure antibodies against the SARS-CoV-2 nucleocapsid protein, thus, may not be of use in determining vaccination responses. Uniquely, the TBS assay detects SARS-CoV-2 antibodies against the trimeric spike protein, which is the predominant immune response produced in response to SARS-CoV-2 vaccination. It is our belief that longitudinal measurement of antibody responses following vaccination should be conducted using multiple assays which measure antibodies against the spike protein, and that this should be done alongside frequent RT-PCR testing. Such experimental design may allow determination of protective antibody levels, as well as cross-comparative evaluation of the performance of several anti-spike serological assays. We envisage this as a multi-national study, analogous to the current SIREN-study. Such studies are urgently required.

Nevertheless, we believe nucleocapsid assays still have a role in sero-epidemiological surveillance. We envisage dual serological testing from use of nucleocapsid and spike assays. This will enable identification of new seroconverts from natural infections and monitoring vaccination responses in niche clinical scenarios such as in poor-vaccine responders or the immunocompromised. It is paramount proficient clinical interpretation is available in sites utilising such diagnostic strategies.

The present study has some limitations. Firstly, our sample sets may under-represent ethnic groups, did not comprise children and did not capture sufficient clinical metadata required for full clinical and analytical result interpretation. We propose a further study to be conducted, where along with participant’s PCR status and clinical symptoms, investigation of these immunoassays to detect responses in asymptomatic and mild cases should be evaluated to deduce the clinical sensitivity of different immunoassays. Whilst limited, the study highlights that the TBS assay showed enhanced sensitivity in detecting mild and asymptomatic antibody responses [9].

## Conclusion

We report the evaluation of three different SARS-CoV-2 antibody assays conducted across two sites. We demonstrated that the Abbott assay showed reduced sensitivity when compared to the Roche and TBS assay, and significantly underestimates seroprevalence. The findings provide evidence that Abbott manufactures threshold should be redefined (0.64 index value) which increased sensitivity (90%) and reduced the incidence of false negatives. The new re-defined threshold produced results that are comparable to both the Roche and TBS assay. Whilst the Abbott IgG nucleocapsid immunoassay has insignificant value in longitudinal serological monitoring due to lack of sensitivity, the TBS anti-spike assay showed enhanced sensitivity in detecting waning antibody responses. This provides evidence that TBS can be utilised as a viable alternative, which meets bespoke clinical and possible geographical needs, where high-throughput assays are unavailable on site.

## Data Availability

Data available upon request

## Acknowledgements

We would like to thank The Binding Site Ltd (Birmingham, UK) for the provision of TBS kits used in this study. We thank Miles Goble (PHU) for his help with setting up the TBS assay on the DS2 platform. We also thank George Baker for his work on retrieving and collecting samples for this study. We thank the University of Manchester for providing remote access of their statistical software packages.

## Authorship Contributions

All authors contributed to the study design. Sample analysis and data analysis was conducted by DM, LT and CW. Critical analysis were performed by AW and MB. The first draft of the manuscript was prepared by DM. All authors commented on previous versions of the manuscript and read and approved the final manuscript version to be published.

## Conflicts of interest/Competing interests (include appropriate disclosures)

All authors declare they have no known conflict of interest or competing interests which could influence the work reported in this paper.

## Availability of data and material (data transparency)

Data available upon request

## Code availability (software application or custom code)

Not applicable.

## Authors’ contributions (optional: please review the submission guidelines from the journal whether statements are mandatory)

Not applicable.

## Consent to participate (include appropriate statements)

All samples were derived after informed consent was gained from volunteers.

## Consent for publication (include appropriate statements)

All authors and volunteers in this study provide consent for publication.

