## Supplemental Table 1 for "Comparative assessment of SARS-CoV-2 serology in healthcare workers with Abbott Architect, Roche Elecsys and The Binding site ELISA immunoassays"

**Supplementary Data**

Table S1. Summary of performance characterises of each immunoassay as provided by the manufacturer.

| Assay and analyser used | Viral target and antibody type | Sample type | Sensitivity (95% CI) on samples taken ≥14 days post-symptom onset/post-RT-PCR positive, [sample numbers] | Specificity (95% CI), number of samples | Manufactures threshold value |
|---|---|---|---|---|---|
| Abbott SARS-CoV-2 Immunoassay, Architect i2000SR | Nucleocapsid protein, IgG | Serum, serum separator tube and plasma. | 100% (95.89-100), [88] | 99.63% (99.05, 99.90), [1070] | Negative: <1.4 Positive: ≥1.4 |
| Roche Elecsys® Anti-SARS-CoV-2, Cobas e 411 | Nucleocapsid protein, IgG | Serum collected using standard sampling tubes. Li-heparin, K2-EDTA and K3-EDTA plasma | 99.5% (97.0-100%), [496] | 99.80% (99.69, 99.89%), [10,453] | Non-reactive: <1.0 Reactive: ≥1.0 |
| The Binding Site Anti IgG/A/M SARS-CoV-2 ELISA, Dynex DS2 | S1/S2 spike protein, Total antibody | Serum, serum separating tube. | 98.6 % (92.6-100), [162] | 98.3 (96.4-99.4), [707]. | Negative: <1.0 Positive: ≥1.0, |
